# Multi-modal investigation of the schizophrenia-associated 3q29 genomic interval reveals global genetic diversity with unique haplotypes and segments that increase the risk for non-allelic homologous recombination

**DOI:** 10.1101/2021.11.10.21266197

**Authors:** Feyza Yilmaz, Umamaheswaran Gurusamy, Trenell J. Mosley, Yulia Mostovoy, Tamim H. Shaikh, Michael E. Zwick, Pui-Yan Kwok, Charles Lee, Jennifer G. Mulle

**Affiliations:** The Jackson Laboratory for Genomic Medicine, Farmington, CT, USA; Cardiovascular Research Institute and Institute for Human Genetics, UCSF School of Medicine, San Francisco, CA, USA; Graduate Program in Genetics and Molecular Biology, Laney Graduate School, Emory University, 201 Dowman Drive, Atlanta, GA, 30322, USA; Department of Pediatrics, Section of Genetics and Metabolism, University of Colorado School of Medicine, Aurora, CO, USA; Department of Genetics, School of Arts and Sciences, Rutgers University, New Brunswick, NJ, USA; Department of Dermatology, UCSF School of Medicine, San Francisco, CA, USA; Precision Medicine Center, The First Affiliated Hospital of Xi’an Jiaotong University, 277 West Yanta Road, Xi’an 710061, Shaanxi, People’s Republic of China; Department of Psychiatry, Robert Wood Johnson Medical School, Rutgers Biomedical and Health Sciences, Rutgers University, New Brunswick, NJ, USA

## Abstract

Chromosomal rearrangements that alter the copy number of dosage-sensitive genes can result in genomic disorders, such as the 3q29 deletion syndrome. At the 3q29 region, non-allelic homologous recombination (NAHR) between paralogous copies of segmental duplications (SDs) leads to a recurrent ∼1.6 Mbp deletion or duplication, causing neurodevelopmental and psychiatric phenotypes. However, risk factors contributing to NAHR at this locus are not well understood. In this study, we used an optical mapping approach to identify structural variations within the 3q29 interval. We identified 18 novel haplotypes among 161 unaffected individuals and used this information to characterize this region in 18 probands with either the 3q29 deletion or 3q29 duplication syndrome. A significant amount of variation in haplotype prevalence was observed between populations. Within probands, we narrowed down the breakpoints to a ∼5 kbp segment within the SD blocks in 89% of the 3q29 deletion and duplication cases studied. Furthermore, all 3q29 deletion and duplication cases could be categorized into one of five distinct classes based on their breakpoints. Contrary to previous findings for other recurrent deletion and duplication loci, there was no evidence for inversions in either parent of the probands mediating the deletion or duplication seen in this syndrome.

## Introduction

Genomic disorders account for a substantial fraction of physical, neurodevelopmental, and psychiatric morbidity ^1–3^. Examples include deletions at the 7q11.23 region known as Williams–Beuren syndrome (WBS, <u>OMIM 194050</u>), deletions at the 22q11.2 locus that give rise to 22q11.2 deletion syndrome, (<u>OMIM 611867</u>), and reciprocal pathogenic deletions, duplications at the proximal 16p11.2 locus (<u>OMIM 611913</u> and <u>OMIM 614671</u>), and the 3q29 deletion syndrome. ^4–9^. The 3q29 deletion syndrome (<u>OMIM 609425</u>) was first identified by Rossi and colleagues in 2001 as a cryptic subtelomeric deletion, and later Willatt and colleagues described an additional six individuals with similar 3q29 deletions ^7,10^. In 2008, reciprocal 3q29 duplications were identified in 19 individuals when analyzing the genomes of 14,698 individuals with idiopathic mental retardation using array comparative genomic hybridization (aCGH) ^11^. The 3q29 deletion syndrome is associated with a >40-fold increased risk for schizophrenia, and patients are also predisposed to developmental delay, intellectual disability, autism spectrum disorder (ASD), anxiety disorders, attention-deficit/hyperactivity disorder (ADHD), congenital heart defects, and additional somatic, neurodevelopmental, and psychiatric phenotypes ^12–14^. Similarly, phenotypes observed in individuals with the reciprocal 3q29 duplication syndrome include developmental delay, speech delay, intellectual disability, ocular and cardiac anomalies, microcephaly, dental anomalies, obesity, seizures, and behavioral similarities to ASD ^11,14–23^. The 3q29 deletion syndrome has an estimated population prevalence of 1:30,000 ^24^, and 3q29 duplication syndrome has a population prevalence of 1:8,000-75,000 ^14^. Although most deletions are *de novo*, a small number of inherited deletions (7%) have been reported ^25^.

The 3q29 deletion is typically *de novo*, yet most patients have the same ∼1.6 Mbp region of chromosome 3 deleted. These observations suggest there are key risk structures flanking the 3q29 interval that predispose to non-allelic homologous recombination (NAHR) and consequently, formation of the deletions and duplications. It has previously been noted that complex regions of the human genome, such as the 3q29 region, can predispose individuals to recurrent deletions or duplications associated with genomic disorders ^26^. Therefore, characterizing the fine structure of these regions among probands, their parents, and unaffected individuals can help us understand the etiology of the genomic disorder as well as whether certain haplotype configurations are at risk for increased frequency of the genomic disorder. In this study, our aim was to determine the fine structure of the 3q29 region in probands with 3q29 deletion or 3q29 duplication syndromes and their parents. We used an optical mapping technique ^27,28^ to identify the haplotype structures and breakpoints in 16 probands with the 3q29 deletion and two probands with the 3q29 duplication syndrome. Additionally, we applied the same approach to identify the haplotype structures in the 3q29 region among 161 unaffected individuals from the 1000 Genomes Project and the California Initiative to Advance Precision Medicine. The results from our study have important implications for the understanding of the molecular etiology of the 3q29 deletion and duplication syndromes.

## Materials and Methods

### Sample collection - 1000 Genomes Project and California Initiative to Advance Precision Medicine cohorts

In this study, we analyzed a ∼2 Mbp region on chromosome 3 (chr3:195,428,934-197,230,596; GRCh38), including three segmental duplication (SD) blocks, denoted as SDA, SDB, and SDC. The two SDs closest to the telomere (SDB and SDC) flank the canonical ∼1.6 Mbp interval deleted in 3q29 deletion syndrome. We investigated this region in 161 unaffected individuals from 26 diverse populations consisting of five super populations, Africans (AFR) (n=37), Americans (AMR) (n=52), East Asians (EAS) (n=22), Europeans (EUR) (n=27), and South Asians (SAS) (n=23), as part of the 1000 Genomes Project (1000GP) and California Initiative to Advance Precision Medicine (CIAPM) (Table S1). CIAPM samples were collected as described previously ^29^. CIAPM samples and 114 unaffected individuals from 1000GP were designated as the University of California San Francisco (UCSF) dataset. Samples from two publicly available datasets, the Human Genome Structural Variation Consortium (HGSVC) and the Human Pangenome Reference Consortium (HPRC), were included in the analyses. HGSVC included Bionano Genomics optical mapping data of three 1000GP samples (HG01573, HG02018, and GM19036), which were previously not studied (TableS1, Web Sources). 44 unaffected individuals from 1000GP had optical mapping data and phased assemblies available through HPRC, two of which (HG00733 and NA19240) were shared between HGSVC and HPRC (Table S1, Web Sources). Additionally, our dataset included optical mapping data of 114 samples that were part of the 1000GP and the CIAPM, three of which (HG00513, HG00732, and NA19239) were shared with HGSVC and not included (Table S1). Cell lines of these additional 1000GP samples were obtained from Coriell and grown in RPMI 1640 media with 15% FBS, supplemented with L-glutamine and penicillin/streptomycin, at 37°C and 5% CO2.

### Sample collection - The 3q29 Project samples

Subjects were recruited from the 3q29 Project registry ^12^ (3q29deletion.org) as previously described ^30^. Inclusion criteria were a validated clinical diagnosis of 3q29 deletion syndrome where the subject’s deletion overlapped the canonical region (chr3:195,725,000– 197,350,000; GRCH37) by ≥ 80%, and willingness and ability to travel to Atlanta, Georgia. Exclusion criteria were any 3q29 deletion with less than 80% overlap with the canonical region and nonfluency in English. 3q29 study subjects underwent deep phenotyping according to an established protocol ^30^. Whole blood drawn at the study visit was used for downstream optical mapping analysis. Optical mapping analysis was completed for 16 probands and parental genomes were analyzed when available. For 10 probands, both biological parents were available for analysis (deletions were confirmed to be *de novo*). Four probands had one parent available for analysis; two probands were singletons (Table S2). Two additional probands with the 3q29 duplication syndrome were also included in the present study; the same protocol was followed for optical mapping analysis. All subjects had genotyping arrays completed for parent-of-origin analysis ^31^.

### High-molecular-weight DNA extraction

Ultra-high-molecular-weight DNA was extracted from cell lines of 1000GP and CIAPM samples (n=114), and from whole blood samples from the 3q29 Project (n=46) according to the Bionano Prep SP Fresh Cells DNA Isolation protocol, revision C (Document #30257), using a Bionano SP Blood & Cell DNA Isolation Kit (catalog #80030). In short, 1.5 million cells were centrifuged and resuspended in a solution containing detergents, proteinase K, and RNase A. DNA was bound to a silica disk, washed, eluted, and homogenized via 1- hour end-over-end rotation at 15 rpm, followed by an overnight rest at room temperature. Isolated DNA was fluorescently tagged at motif CTTAAG by the enzyme DLE-1 and counter-stained using a Bionano PrepTM DNA Labeling Kit – Direct Label and Stain (catalog #8005) according to the Bionano Prep DLS Protocol, revision F (Document #30206). A total of 750 ng of purified genomic DNA was labeled by incubating with DL-Green dye and DLE-1 Enzyme in DLE-1 Buffer for 2 hours at 37°C, followed by heat inactivation of the enzyme for 20 minutes at 70°C. The labeled DNA was treated with Proteinase K at 50°C for 1 hour, and excess DL-Green dye was removed by membrane adsorption. The DNA was stored at 4°C overnight to facilitate DNA homogenization and then quantified using a Qubit dsDNA HS Assay Kit (Molecular Probes/Life Technologies). The labeled DNA was stained with an intercalating dye and left to stand at room temperature for at least 2 hours before loading onto a Bionano Chip. The DNA was loaded onto the Bionano Genomics Saphyr Chip and linearized and visualized by the Saphyr systems. Data collection was performed using Saphyr 2nd generation instruments (Part #60325) and Instrument Control Software (ICS) version 4.9.19316.1. The DNA backbone length and locations of fluorescent labels along each molecule were detected using the Saphyr system software.

### De novo assembly of optical genome maps

Single-molecule genome maps of 1000GP, CIAPM, and The 3q29 Project samples were assembled *de novo* into genome maps using the assembly pipeline Bionano Solve v3.5, developed by Bionano Genomics with default settings ^32^ as described previously ^33^. In short, a pairwise comparison of DNA molecules (min 250 kbp) was generated to produce the initial consensus genome maps. During an extension step, molecules were aligned to genome maps, and maps were extended based on the molecules aligning past the map ends. Overlapping genome maps were then merged. Extension and merge steps were repeated five times before a final refinement of the genome maps. Clusters of molecules aligned to genome maps with unaligned ends >30 kbp in the extension step were re-assembled to identify all alleles. To identify alternate alleles with smaller size differences from the assembled allele, clusters of molecules aligned to genome maps were detected with internal alignment gaps of size <50 kbp, in which case the genome maps were converted into two haplotype maps. The final genome maps were aligned to the reference genome GRCh38.

### Single-molecule analysis pipeline for optical genome maps

In the 3q29 SD blocks, SDA, SDB, and SDC, we defined segments according to characteristic banding patterns of Bionano Genomics optical mapping labels. We color-coded these segments and displayed them according to assigned colors. The order of the segments are: SDA-26 kbp (magenta), SDA-5kbp (blue), SDA-11 kbp (yellow), SDA-4 kbp (red), ∼33 kbp (maroon, SDA and unique region), ∼15 kbp (orange, unique region), ∼124 kbp (green, unique region), SDB-5kbp 1st copy (blue), SDB-15kbp (magenta partial copy), SDB-4 kbp (red), SDB-11kbp (yellow), SDB-5kbp 2nd copy (blue), SDC-11 kbp (yellow), SDC-5 kbp (blue), and SDC-15 kbp (magenta partial copy) (Table S3). Structural variations and haplotypes at the 3q29 locus were analyzed and single molecules were evaluated in 161 unaffected individuals from 26 diverse populations from the 1000GP and CIAPM, and 46 samples from the 3q29 Project using the Optical Maps to Genotype Structural Variation (OMGenSV) package as described previously ^34,35^. To identify 3q29 haplotypes in samples from 114 unaffected individuals which were part of 1000GP and CIAPM, the assembled contigs were visualized using the “anchor” mode in OMView from the OMTools package ^36^, and Bionano Access was used for HGSVC (n=3), HPRC (n=44), and the 3q29 Project (n=46) samples. Haplotypes were manually identified from these visualizations, and corresponding consensus map (cmap) files were constructed for each haplotype if the contig in the 3q29 region was not contiguous, including at least 500 kbp of the unique flanking region where applicable. When 3q29 SDs contained multiple SVs and were too long to analyze from end to end with single molecules (>350 kbp), molecules were subdivided into groups that were anchored in the proximal or distal unique regions of 3q29 SDA and SDB. For each haplotype, the corresponding cmaps were compiled into a single file and used as a reference input file for the OMGenSV pipeline, along with local molecules from each sample and a set of “critical regions’’ (SDA:195,578,485-195,817,578 Mbp; SDB:195,804,017-196,073,500 Mbp; SDC:197,557,633-197,743,251 Mbp; GRCh38) defining the areas on each cmap that molecules need to span to support the presence of that haplotype in the sample. The 3q29 Project sample cohort haplotypes were identified by using 1000GP and CIAPM individuals’ 3q29 haplotype maps as a reference, and molecule support was confirmed by following the steps described above. Images of molecule support were obtained by using OMTools and Bionano Access (Data and Code Availability). Fisher’s exact test was used to test the significance of the 3q29 haplotypes in R Studio (version 1.4.1717). 100,000 replicates were used for the Monte Carlo test. The expected value for each haplotype was calculated by the following command:

~~~
*tab.exp <- round(chisq.test(data)$expected)*
~~~

Then we used expected values for Fisher’s exact test:

~~~
fisher.test(data, simul=T, B =100000)$p.value
~~~

Association plots were generated with the following command:

~~~
tab.sel <- as.matrix(data[freq$Total >5,])
assocplot(tab.sel, col=c(“red”,“green”))
~~~

Finally, we annotated all haplotypes detected in our study with GENCODE v37 to identify overlapping genomic elements, including protein-coding genes and noncoding RNAs.

### Breakpoint mapping and trio analysis of the 3q29 Project samples

For each proband (n=18, 16 probands with the 3q29 deletion and two probands with the 3q29 duplication syndrome), *de novo* assembled contigs were aligned to the GRCh38 human genome reference assembly, and the 3q29 region was manually inspected to identify deletion and duplication breakpoints. Once identified, the underlying single molecules were examined to verify that deletion and duplication breakpoints were well supported by single molecules (n ≥ 3 molecules). Then, the sequence identity of approximate breakpoints and the corresponding sequence in GRCh38 was determined by the NCBI *blastn* tool (BLASTN 2.9.0+) with the following command:

~~~
blastn -query $file1 -subject $file2 -num_alignments 5 –
num_descriptions 5 -out $alignmentoutputfile
~~~

Only “the best alignment” was included in the final output file. Haplotypes of the parents were identified as described above. Finally, the parents transmitting the intact chromosome and deletion parent of origin were identified by visual pairwise comparison of the haplotypes using Bionano Access.

### Validation using orthogonal data

Publicly available optical mapping data and phased assemblies of 44 unaffected individuals as part of the 1000GP from the HPRC were analyzed to confirm the 3q29 haplotypes which were identified by using Bionano Genomics optical mapping data. First, HPRC phased assembly fasta files were converted to *in silico* maps and haplotypes were detected as described above. Then a visual pairwise comparison of optical mapping and phased assembly 3q29 haplotypes for each sample was performed by using Bionano Access (Figure S1-S4). All HPRC phased assemblies (n=44) except the paternal haplotype of HG01928 supported the haplotypes identified in optical mapping data (Figure S5).

## Results

The 3q29 region contains three segmental duplication (SD) blocks within a ∼2 Mbp region, SDA (∼73 kbp), SDB (∼64 kbp), and SDC (∼41 kbp) (GRCh38) (Figure 1, Table S3). Colored arrows depicting the 3q29 segments and their orientations were determined based on the labeling pattern from the optical mapping data and the sequence identity between 3q29 SDs (Table S4) as described in the Materials and Methods. In the GRCh38 human genome reference assembly we identified three copies of ∼26 kbp segment (magenta), four copies of ∼5 kbp (blue), three copies of ∼11 kbp (yellow), two copies of ∼4 kbp (red), one copy of ∼33 kbp (maroon), one copy of ∼15 kbp (orange), and one copy of ∼124 kbp (green) segments (Figure 1B).

**Figure 1:**
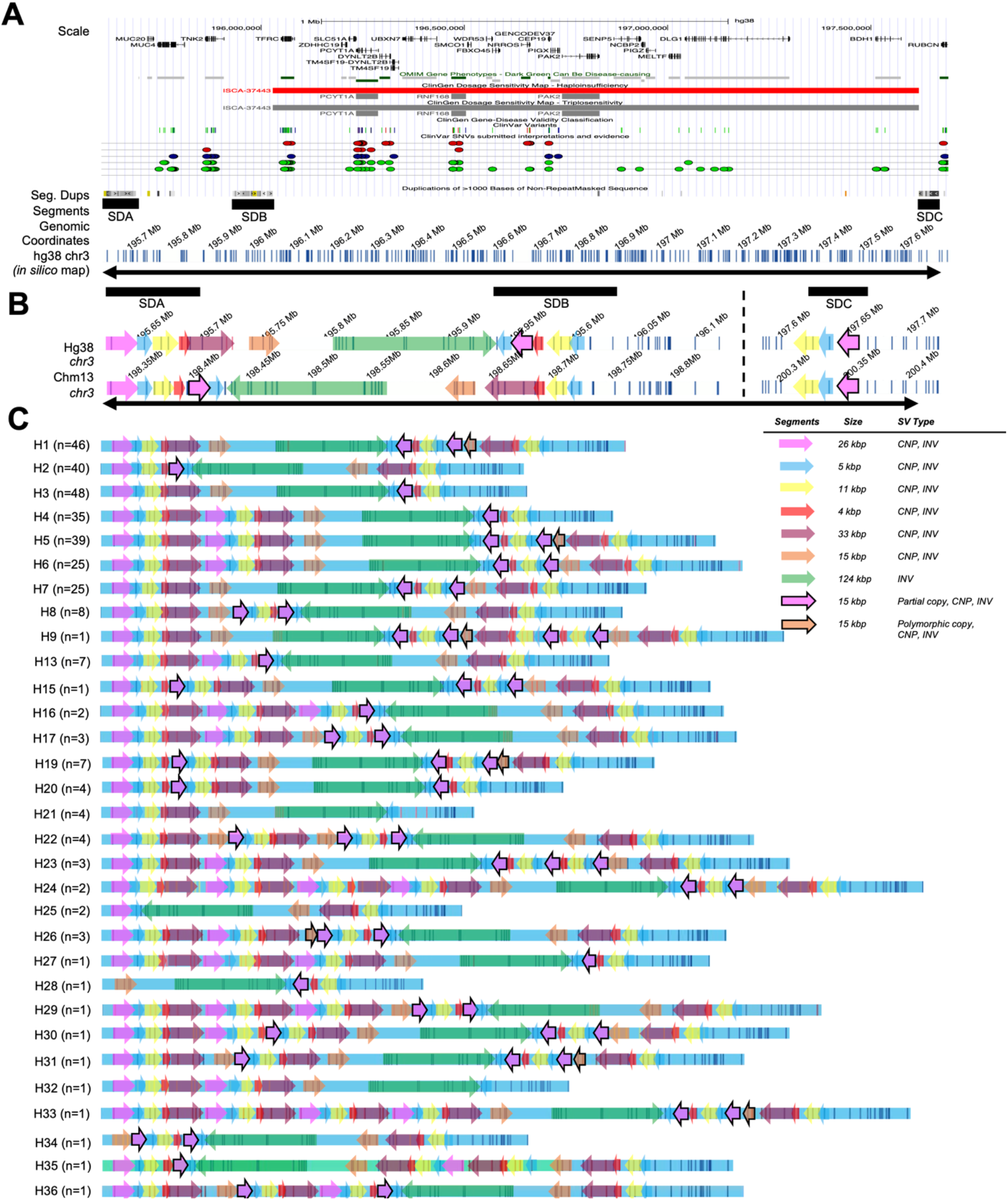
The segment structure of GRCh38 3q29 region and 3q29 haplotypes identified in this study. (A) 3q29 genomic locus with SDA, SDB, SDC, OMIM Gene Phenotypes, ClinGen Dosage Sensitivity Map - Haploinsufficiency, ClinGen Dosage Sensitivity Map - Triplosensitivity, ClinGen Gene-Disease Validity Classification, ClinVar Variants, ClinVar SNVs, and Duplications of > 1000 Bases of Non-RepeatMasked Sequence represented as Tracks. The 3q29 GRCh38 in silico map is represented as the last track. (B) 3q29 segments of GRCh38 and T2T/chm13 with SDA, SDB, and SDC represented as black boxes on top and genomic segments as colored arrows overlaid on the in silico maps (white background with vertical blue lines). Dotted line: the unique region between SDB and SDC, 196.1-197.6Mbp. (C) The structure and prevalence of 13 previously reported and identified among our samples (H1-H9, H13, H15-H17) and 18 novel 3q29 haplotypes (H19-H36) of 3q29 SDA and SDB. Black arrows in panels A and B represent the region included in the analyses. Novel haplotypes were ordered by prevalence in the population. Each colored arrow, magenta, blue, yellow, maroon, orange, green, represents 3q29 segments. CNP: copy number polymorphism, INV: inversion.

### 3q29 haplotypes in unaffected individuals from 1000GP and CIAPM

In this study, we used *de novo* assembled optical genome maps having an average effective coverage of 102.4 X and an average N50 of 301.96 kbp. We identified a total of 36 different haplotypes for the 3q29 region (195,605,705 - 197,706,750; GRCh38) among 161 individuals from the 1000GP and CIAPM cohort. Within the SDC region, no variation was observed across samples. Therefore, all haplotypes were based on the variation observed between SDA and SDB. These 161 individuals were from 26 diverse populations representing five super populations: AFR, AMR, EAS, EUR, and SAS (Table S2). The haplotype size of the 3q29 region ranged from ∼287 kbp (H28, HG03863-SAS) to 859 kbp (H24, BC02901-AMR, and BC03702-EUR) (Figure 1, Table S5). The smallest haplotype, H28, is missing the proximal ∼440 kbp and the distal ∼132.5 kbp of the largest haplotype H24 and lacks five copies of the following segments: ∼26 kbp, ∼5 kbp, ∼11 kb, and ∼33 kbp (Figure S6). Of 18 previously described haplotypes, we identified 13 haplotypes in our samples; H10, H11, H12, H14, and H18 from Ebert and colleagues ^33^ were not observed. We detected 18 novel haplotypes, H19-H36. The most common three haplotypes among all 161 unaffected individuals were H3 (14.91%), H1 (14.29%), and H2 (12.42%). The H3 haplotype (14.91%) represents the GRCh38 human genome reference assembly haplotype for the 3q29 region, and the H2 (12.42%) haplotype was described in the most recent human genome assembly as part of the Telomere-to-Telomere (T2T) Consortium ^37^. We also detected 14 haplotypes that were each observed once among all 161 genomes (i.e., 322 chromosomes) examined and therefore referred to as “singletons”. To assess the accuracy of the haplotypes identified by optical mapping data, we overlaid long-read sequencing-based phased assemblies (HPRC dataset, n=44 (Table S5)) to the optical mapping-based haplotypes. 20 haplotypes (11 previously identified and 9 novel) assessed in this manner were confirmed by long-read phased assemblies. One discrepancy was detected between the optical mapping and phased assembly haplotype structures, for one chromosome from sample HG01928. The phased assembly suggested that haplotypes were H1/H5, and the optical mapping data suggested that haplotypes were H4/H5. Single DNA molecules (>10) from the optical mapping data were anchored to the proximal and distal unique region and span at least 90% of the 3q29 region and confirmed the presence of the H4 haplotype in HG01928 (Figure S5). Our data suggest that the phased assembly for this individual may have been incorrectly assembled.

We found significant differences in haplotype frequency between populations (Figure 2, *p-value = 0.000001 Fisher’s exact test*). One of the most common haplotypes, H3, was observed at ≥ 7% frequency (≥ 14% in four populations) in all five super populations (Figure 2A). In the AFR population, three haplotypes account for 48% of the haplotype pool: H2 (22%), which was significantly enriched compared to other haplotypes and all other populations (*p-value = 0.009, Fisher’s exact test*), H3 (15%), and H8 (11%) (Figure 2B). Interestingly, the H8 haplotype, which contains the second-largest inversion observed in the 3q29 region (∼289 kbp), was observed only in the AFR population (Figure 2B, *p-value = 5.73972e-06, Fisher’s exact test*). Similarly, H19 haplotype was enriched in the AFR population (p-value = *5.73972e-06*, Fisher’s exact test) Furthermore, the H5 haplotype was enriched in the EAS population (41%, p-value = 7.731026e-08, Fisher’s exact test compared to other haplotypes and all other populations). The H6 haplotype was enriched in the EUR population (19%, p-value = 0.0002, Fisher’s exact test compared to all other populations and haplotypes). The H1 haplotype was enriched in the SAS population (26%, p-value = 0.002, compared to other haplotypes and all other populations based on the Fisher’s exact test). The SVs differ from one haplotype to the other, often overlapping protein-coding genes (e.g., *MUC4* and *MUC20*), pseudogenes (e.g., *SDHAP1* and *SDHAP2*), and lincRNAs (e.g., lncRNA MUC20-OT1).

**Figure 2:**
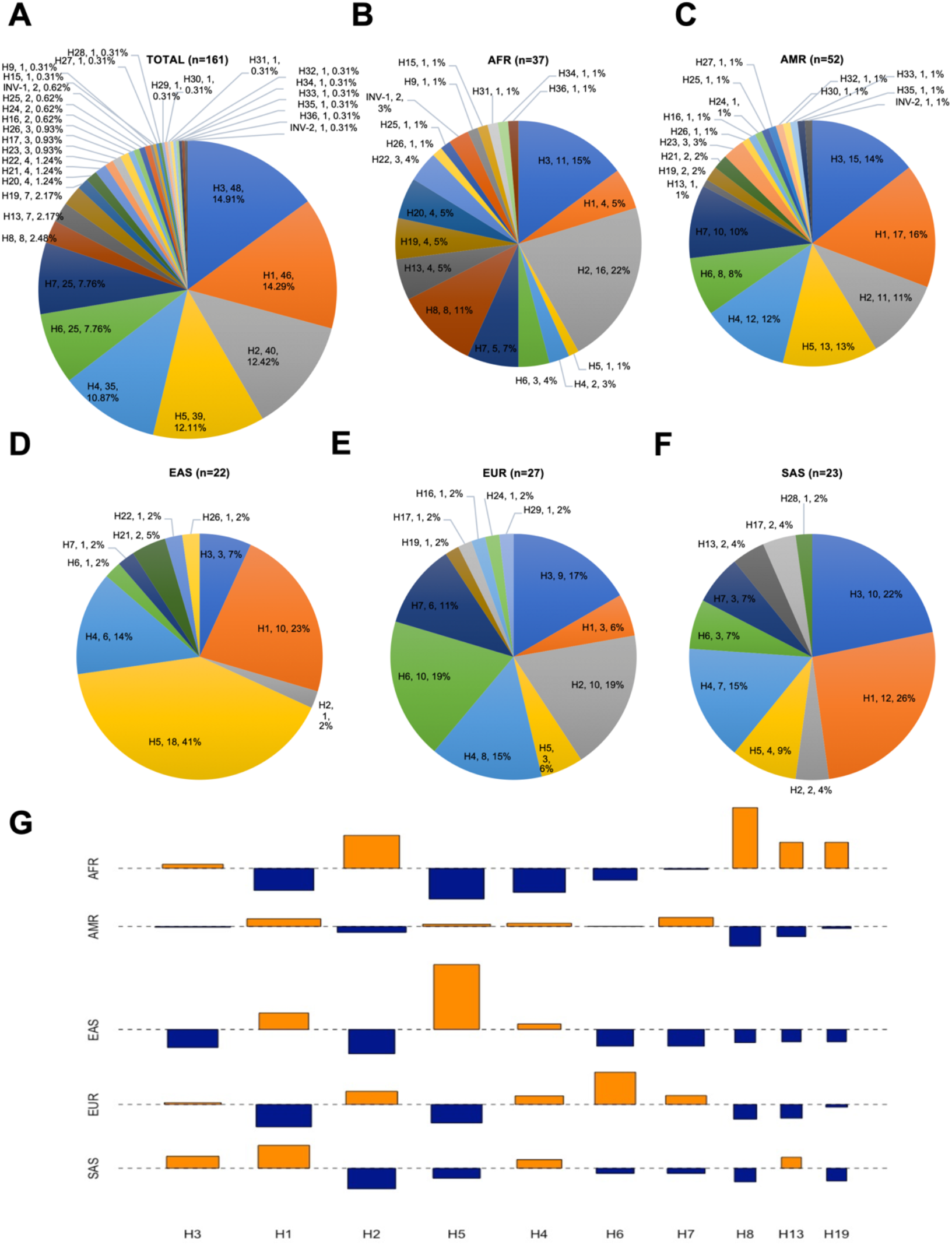
Prevalence of the 3q29 haplotypes represented in five major populations. We identified 3q29 haplotypes in each sample and then calculated the prevalence of 3q29 haplotypes in five major populations. (A) The distribution of haplotypes identified in total. (B) the distribution of haplotypes in the AFR population, (C) the distribution of haplotypes in the AMR population. (D) the distribution of haplotypes in the EAS population. (E) the distribution of haplotypes in the EUR population, (F) The distribution of haplotypes in the SAS population. AFR - African, AMR - American, EAS - East Asian, EUR - European, SAS - South Asian. Each color in the pie charts represents a distinct 3q29 haplotype. The order in the labels in pie charts represents haplotype ID, count, prevalence. Singleton haplotypes are H9, H15, H27-H36, and INV-2. n represents the number of samples. (G) - Cohen-Friendly association plot depicting the relationship between haplotypes and populations. Each cell has a rectangle with height proportional to the difference between observed and expected (normalized to expected) and width proportional to the square root of expected counts so that the area of the rectangle is proportional to the difference in observed and expected counts. Expected values were calculated as described in the Materials and Methods. If the observed count is greater than expected, the rectangle rises above the baseline and is colored in red.

### Inversions among the 3q29 haplotypes

Among the 36 haplotypes identified from unaffected samples, we detected three different types of inversions that were > 100 kbp in size: two large inversions (∼ 2.03 Mbp and ∼2.13 Mbp) between SDA and SDC, and a smaller inversion (∼289 kbp) between SDA and SDB which was observed on twelve distinct haplotypes (Figure 3, Figure S3). Additionally, we observed inversions of each 3q29 segment in distinct haplotypes. The most prevalent inversion was a ∼289 kbp inversion which was observed at a frequency > 10% in all five super populations on the following haplotypes: H2, H8, H13, H16, H17, H22, H25, H26, H29, H34, H35, and H36 (Figure 3A, Figure S7). This previously reported inversion lies in a unique region between the 3q29 SDA and SDB blocks and was observed in seven novel and five previously identified haplotypes in 69 unaffected individuals (76 chromosomes, Figure S3, Table S6) ^38,39^. The two larger >2 Mbp inversions between 3q29 SDA and SDC (Figure 3, Figure S8) were identified among three unaffected individuals. The ∼2.13 Mbp inversion (INV-1) (Figure 3B) was observed in HG03470-AFR and potentially arose on haplotype H8, and the ∼2.03 Mbp inversion (INV-2) (Figure 3C) was observed in two individuals, GM19984-AFR and BC04902-AMR, and potentially arose on H20 haplotype. The ∼2.03 Mbp and ∼2.13 Mbp inversions are similar to the ∼2 Mbp inversion described previously by Antonacci and colleagues (Figure 3D) ^40^. However, the breakpoints of the ∼2 Mbp inversion were localized to a 15.5 kbp region (chr3:196,868,578 – 196,884,133 and chr3:198,832,975 – 198,848,521; hg17/UCSC 2004), and a more precise definition was not provided because the proximal breakpoint overlapped with an assembly gap in the hg17/UCSC 2004 human genome reference assembly. In this study, we have found that breakpoints for all inversions clustered to a ∼5 kbp (blue) paralogous SD copies within SDA and SDC. These paralogous copies share ∼98% sequence identity which suggests that NAHR is the mechanism causing these inversions.

**Figure 3:**
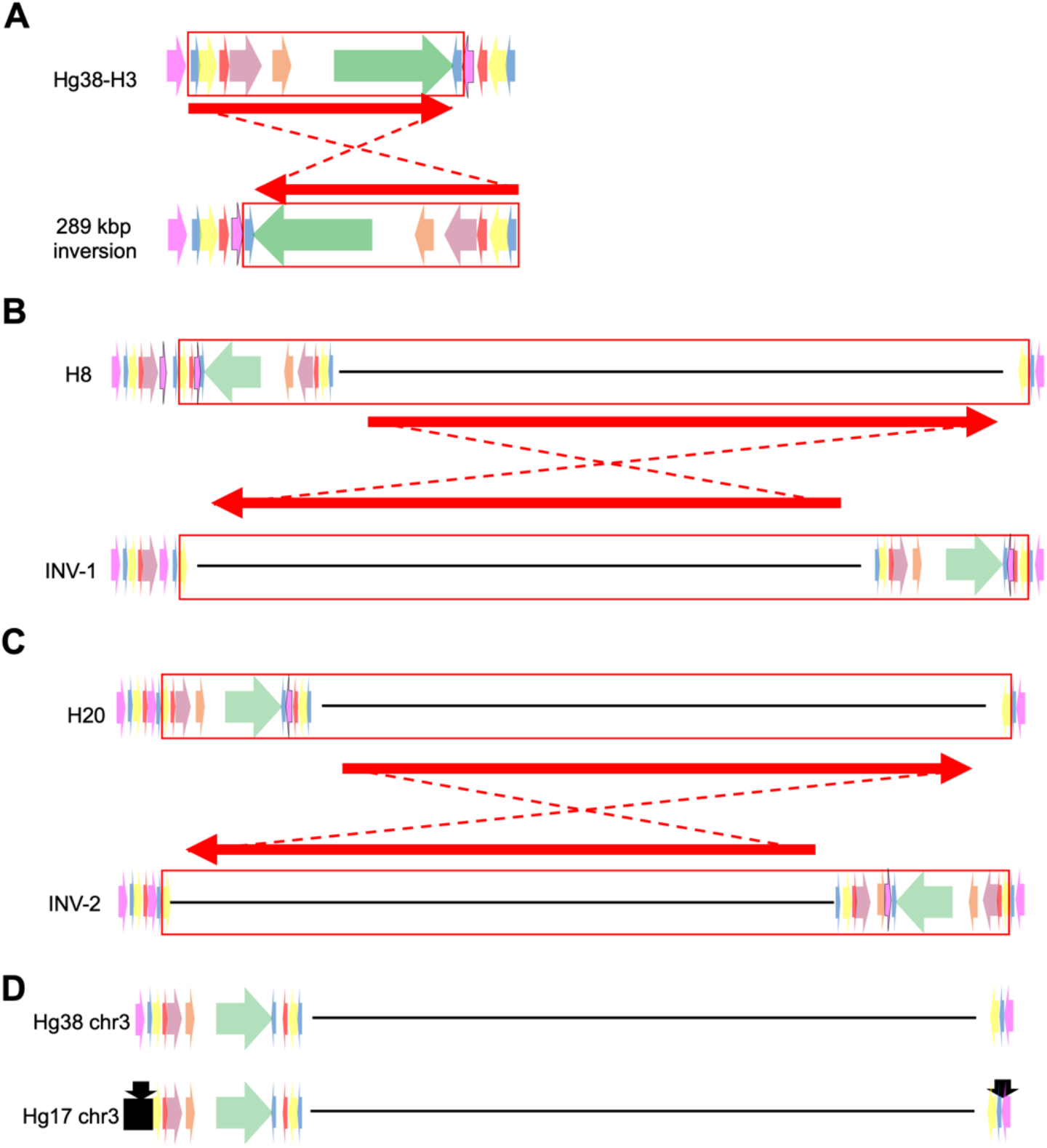
Inversions > 100 kbp identified from the 1000GP and CIAPM samples. (A) Hg38/GRCh38 human genome reference assembly presented at the top and the 289 kbp inversion, including the 5, 11, 4, 33, 15, and 124 kbp segments, represented at the bottom. (B) - Upper rectangle represents the H8 haplotype of the 3q29 region. The rectangle at the bottom represents the structure of INV-1. (C) - Upper rectangle represents the H20 haplotype of the 3q29 region. The rectangle at the bottom represents the structure of INV-2. (D) - Representation of Hg38/GRCh38 and Hg17/NCBI35 reference assemblies. Black arrows in Hg17/NCBI35 represent inversion breakpoints identified by Antonacci and colleagues. Colored arrows in each panel represent 3q29 segments, red rectangles indicate the region involved in the inversion, red arrows represent the orientation of the region before and after the inversion and dashed lines show the inversion.

### 3q29 probands and parental haplotypes

3q29 deletion syndrome is associated with a >40-fold increased risk for schizophrenia and can also be associated with neurodevelopmental disorders. Therefore, characterizing the fine structure of the 3q29 region in probands and their parents is critical to gain a better understanding of disease etiology. In this study, we analyzed 16 probands with the 3q29 deletion and two probands with the 3q29 duplication syndrome from the 3q29 Project using Bionano Genomics optical mapping technology (Figure 4A). The average effective coverage of assembly was 102.4 X and the average molecule N50 was 301.96 kbp (Table S2). The recurrent ∼1.6 Mbp deletion in the 3q29 deletion syndrome probands results from one break in SDB and one break in SDC ^7,10,11^. The unique genomic sequence residing between SDA and SDB is typically intact, not deleted, in the 3q29 deletion syndrome probands. We detected the recurrent ∼1.6 Mbp deletion in 15 probands. In one proband we detected a smaller ∼1.2 Mbp deletion lying completely within the ∼1.6 Mbp 3q29 deletion region (Figure 4A, Family 18). This smaller deletion contains 19 of the 21 genes completely within its breakpoints. The centromeric breakpoint falls within the *TFRC* gene, deleting the promoter and the first four exons. On the telomeric side, the *BDH1* gene is intact. There is no discernible difference in phenotypic severity in this proband, despite their smaller deletion. None of the 15 deletions (∼1.6 Mbp (n=14) and ∼1.2 Mbp (n=1)) were seen in either of the parents and were therefore considered to be *de novo* events. One proband inherited the deletion (∼1.6 Mbp) from his father (Family 20).

**Figure 4:**
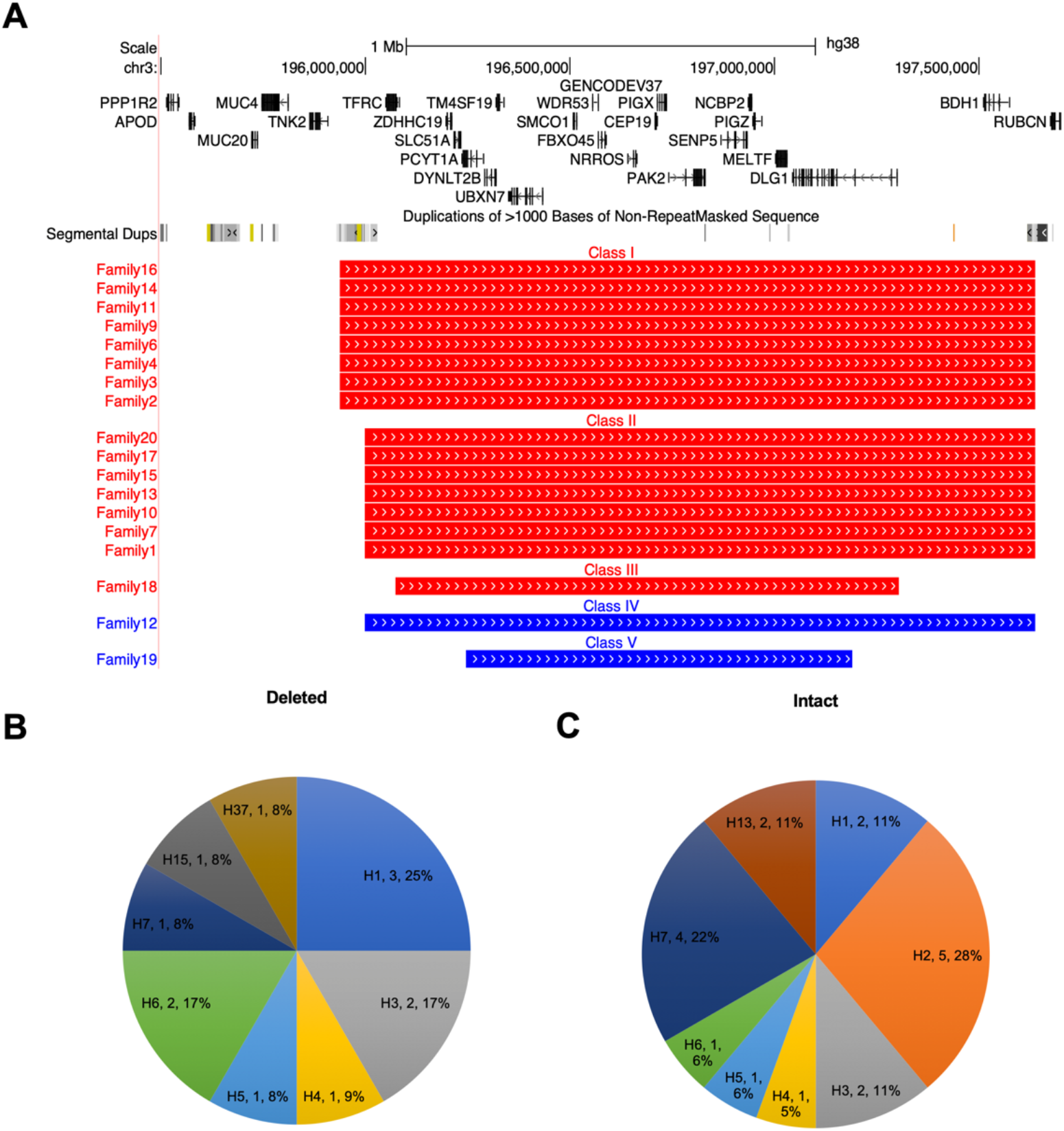
3q29 Deletions and duplications reported among the studied probands. (A) Genes and duplications of > 1000 Bases of Non-Repeat Masked Sequence represented as Tracks overlapping the 3q29 region (top). Red lines represent deletion, and blue lines duplication cases analyzed in this study (bottom). (B) Distribution of the 3q29 haplotypes observed in deleted chromosomes in probands. (C) Distribution of the 3q29 haplotypes observed in intact chromosomes in probands. The order in the labels in pie charts represents haplotype IDs, count, and prevalence.

To investigate whether certain haplotypes predispose to the 3q29 deletion or duplication syndrome, we characterized the 3q29 region, determined the haplotype in probands (Figure 4B-C) and available parents, and finally determined which chromosomes were inherited by the probands. For ten probands, DNA from both biological parents was available (Table S7). For two probands, DNA from only one parent was available (Table S7). For four probands, DNA was not available from either of the parents (Table S7). When possible, we determined on which inherited haplotype the deletion occurred, and which chromosome was inherited in an intact state for each proband. Interestingly, we identified a novel haplotype, H37, in one father (Family 18), who was the parent of origin of the deletion observed in the proband (Figure S9). However, we did not observe an enrichment of 3q29 deletions or duplications with any particular haplotype.

### Defining five breakpoint classes for the 3q29 deletion and duplication syndromes

While it has been known that breakpoints for 3q29 deletions and duplications generally occur within two SD blocks (SDB and SDC), the breakpoints have not been precisely mapped. Among the 16 probands with the 3q29 deletions, we identified three deletion classes (Table S8). The Class I deletion breakpoints (∼1.69 Mbp, 195.94-197.64 Mbp; GRCh38) were observed in eight probands, with the proximal breakpoint localized to the first ∼5 kbp segment (blue, Figure 5A) in SDB, and the distal to a ∼5 kbp paralogous copy in SDC (blue, Figure 5A). Although optical mapping data indicated that the deletion breakpoints localized to the ∼5 kbp segments, the possibility of breakpoint localization to the ∼15 kbp segments (magenta, partial copies of SDA-26 kbp segment) in SDB and SDC cannot be excluded. The optical mapping resolution in this ∼15 kbp was insufficient (only one label resides within ∼26 kbp segment and zero labels in the ∼15 kbp segment, which is a partial copy of ∼26 kbp segment), therefore sequence-level resolution is required to determine whether paralogous copies of ∼5 kbp or ∼15 kbp cause NAHR in deletions classified as Class I. The Class II deletions (∼1.63 Mbp, 195.99-197.64 Mbp; GRCh38) were observed in seven probands and had the proximal breakpoint localized to the second ∼5 kbp segment in SDB and the distal breakpoint localized to the same ∼5 kbp segment in SDC (Figure 5B). The Class III deletion (∼1.23 Mbp, 196.01-197.3 Mbp; GRCh38) was observed in only one proband, and the proximal and distal breakpoints did not overlap with any SDs (Figure 5C). Sequence analysis of the ∼5 kbp segments in SDB and SDC in the GRCh38 reference genome showed a high sequence identity to each other (≥ 98%) suggesting that these paralogous copies can facilitate NAHR-mediated 3q29 deletions and duplications (Figure 5D-G, Table S7). On the other hand, the proximal and distal deletion breakpoints in Class III were not localized to SDs, sequence similarity was not detected between breakpoints which suggests that a mechanism other than NAHR has caused this deletion.

**Figure 5:**
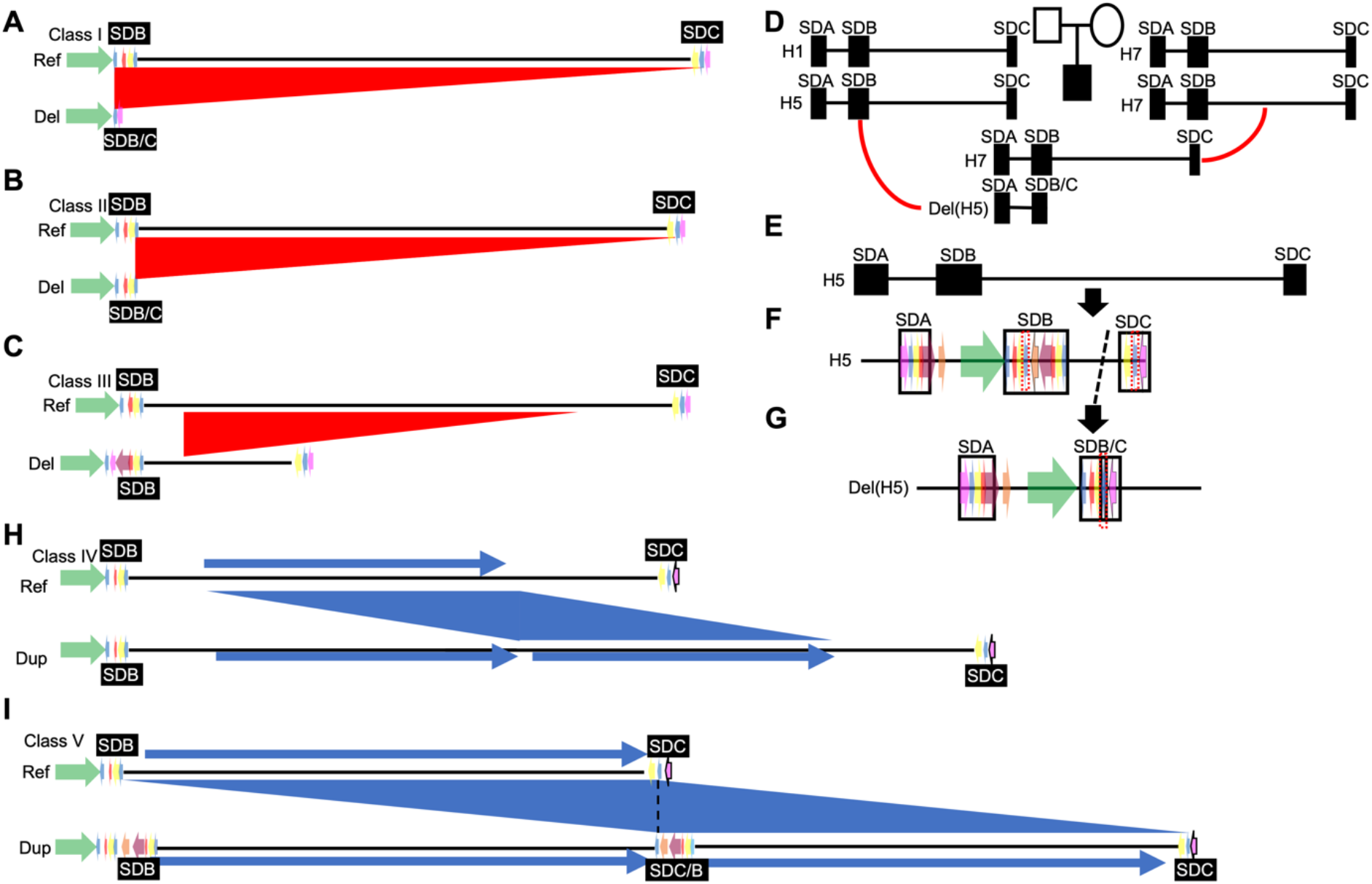
Deletion and duplication breakpoint classes identified in our study and depiction of NAHR-mediated deletion in Family 7 trio from the 3q29 Project. (A-C) Schematic representation of 3q29 deletion breakpoint classes. The red triangles represent the deletion in each class. (D) Pedigree of Family 7, black rectangles with vertical dark lines represent genomes of the father, mother, and proband, red curved lines show the parent of origin and the transmitting parent, and black rectangles represent the 3q29 SDs. (E) Father’s parent of origin chromosome, haplotype H5. (F) The 3q29 SDs, SDA-B-C, and the proposed NAHR mechanism. (G) Deleted chromosome occurring as a result of NAHR. Red dashed boxes indicate the proposed segments to mediate NAHR and result in 3q29 deletion in the proband. (H-I) Duplication breakpoint classes from Families 12 and 19, respectively. Black dotted line represents the duplication breakpoint. Blue rectangle: duplicated 3q29 region with the blue shaded area representing the duplication.

Additionally, we analyzed two probands with duplications, ranging in size from ∼942 kbp (∼196.25-197.2 Mbp; GRCh38) to ∼1.6 Mbp (∼196-197.6 Mbp; GRCh38). We categorized these duplications as Class IV and Class V, respectively (Figure 5H-I). In Class IV, the proximal and distal breakpoints localized to a unique region between SDB and SDC (Figure 5H) suggesting that a mechanism other than NAHR has caused the duplication. The Class V duplication appears to be a reciprocal duplication of the recurrent ∼1.6 Mbp 3q29 Class II deletion (Figure 5I). The proximal breakpoint in Class V localized to the second ∼5 kbp segment (blue) in SDB and the distal breakpoint localized to the paralogous ∼5 kbp segment (blue) in SDC (Figure 5I). Therefore, this duplication was likely caused by NAHR.

Finally, we investigated 3q29 haplotypes and phenotypes to evaluate whether there was a correlation between haplotypes and phenotypes of 3q29 deletion and duplication probands, however, no significant associations were identified (Table S9).

## Discussion

The 3q29 deletion and duplication syndromes are rare genomic disorders. In the recurrent 3q29 deletion and duplication cases, breakpoints occur within two SD blocks which themselves are comprised of different SDs with > 98% sequence identity prone to NAHR ^11,41,42^. Since these SD blocks have a complex structure containing paralogous copies of SDs with high sequence identity, it is often difficult to accurately identify where the breakpoints occurred. Techniques such as short-read sequencing have limited ability to resolve the breakpoint locations within repetitive genomic segments such as SDs, due to the high sequence identity and lengths of the paralogous copies ^43^. With the availability of complementary genomic technologies, including long-read sequencing and optical mapping, we are now able to interrogate these regions and to identify breakpoint locations more precisely ^34,35^.

In this study, we first *de novo* assembled the 3q29 region in 161 unaffected individuals from the 1000GP and CIAPM cohorts to identify distinct haplotypes. Ebert and colleagues ^33^ previously reported 18 haplotypes in 30 unaffected individuals from the 1000GP. We have now identified an additional 18 novel haplotypes in unaffected individuals using optical mapping technology bringing the total number of known haplotypes for this region to 36. Six haplotypes are either population-specific or have low frequency in other populations. For instance, the H8 haplotype was enriched in 11% of the AFR population, the H5 haplotype was enriched in 41% of the EAS population, the H6 haplotype was enriched in 19% of the EUR population, and the H1 haplotype was enriched in 26% of the SAS population. In summary, we have conducted the largest and most detailed investigation of 3q29 haplotypes in 161 unaffected individuals from 26 worldwide populations to date and report significant differences in haplotype frequencies between the studied populations. Furthermore, the 3q29 haplotypes contain inversions and copy number changes of 3q29 segments which overlap with protein-coding genes, pseudogenes, and non-coding RNAs, such as *MUC20* and *MUC4*, lncRNA MUC20-OT1, *SDHAP1*, *SDHAP2*, *SMBD1P*, and miRNAs. A recent study showed that lncRNA *SDHAP1* upregulated the expression of *EIF4G2* by reducing miR-4465 levels in ovarian cancer cells ^44^ which suggests that the pseudogenes may regulate gene expression through microRNAs ^45^. Therefore, additional copies of the pseudogenes may impact patients’ phenotypes by regulating the protein-coding genes in the 3q29 region. However, further studies need to be conducted to investigate the function of the pseudogenes in the etiology of 3q29 deletion and duplication syndromes.

We analyzed the 3q29 haplotype structures in 16 probands with 3q29 deletion syndrome to determine the breakpoint locations more precisely in probands and available parents, and to identify the molecular mechanisms responsible for the deletions. In 15 of 16 probands, the deletion breakpoints overlapped paralogous copies of ∼5 kbp SD segments (blue) within SDB and SDC, located in the same orientation. High sequence identity (> 98%) between the ∼5 kbp segments (blue) suggests that an NAHR mechanism caused the deletions in these probands, consistent with previous findings ^11^. However, in Class I, the possibility of breakpoint localization to the ∼15 kbp segments (magenta, partial copies of SDA-26 kbp segment) in SDB and SDC cannot be excluded since these segments are adjacent (in the distal region) to the ∼5 kbp segment (blue). The sequence-level resolution is required to overcome the optical mapping resolution insufficiency at the ∼15 kbp segment and to determine whether paralogous copies of ∼5 kbp or ∼15 kbp cause NAHR in deletions classified as Class I. The deletion breakpoints in Class II were localized to the paralogous copies of ∼5 kbp segments and the deletion breakpoints for one proband, Class III, did not occur within any SD blocks in the region suggesting that non-homologous end joining (NHEJ) or replication-based mechanism might be responsible ^46^. We also analyzed two individuals with the 3q29 duplication syndrome. One individual carried the common recurrent ∼1.6 Mbp duplication that appears to be the reciprocal duplication of the recurrent ∼1.6 Mbp 3q29 deletion, whereas the other ∼1.2 Mbp 3q29 deletion was not recurrent. Based on the deletion and duplication breakpoints, we have defined three deletion and two duplication breakpoint classes for the 3q29 Project patient cohort.

Inversions have previously been hypothesized to be a risk factor for some *de novo* deletions and duplications ^47^. It is thought that inversions can interfere with synapsis during meiosis, potentially causing DNA loops that are susceptible to misalignment and/or breakage ^2^. For example, at the Williams-Beuren syndrome locus, there is an enrichment of inversions in the parents where the *de novo* deletion arises ^48,49^. However, in 2003, a group studying the 22q11.2 deletion syndrome showed that none of the parents of 18 probands with the 22q11.2 deletion syndrome carried an inversion in this region ^50^. Two inversions, ∼289 kbp, and ∼2 Mbp, have previously been reported at the 3q29 locus ^38–40^. The breakpoints in these inversions were localized to inversely oriented paralogous copies of 3q29 segments between SDA and SDB, and SDA and SDC, which could mediate NAHR. Among the 22 parents analyzed in the current study, six were found to carry the ∼289 kbp inversion, and three were inherited (H2: n=2, H13: n=1) by the proband in an intact state. None of the parents carried the larger ∼2 Mbp inversion occurring between SDA and SDC. While the sample size is relatively small, our data provide no evidence for an inversion predisposing to deletion or duplication at the 3q29 locus.

One limitation of our study is the lack of resolution of deletion and duplication breakpoints at the sequence level. Nevertheless, with optical mapping, we refined the deletion and duplication breakpoints to ∼5 kbp segments. Other technologies, such as long reads from Oxford Nanopore Technology (ONT) or Pacific Biosciences (PacBio) may be able to provide nucleotide resolution breakpoints for these deletions and duplications. Unfortunately, the available DNA samples from the 3q29 Project are not appropriate for ONT or PacBio High Fidelity (HiFi) long-read sequencing since these require large amounts of high molecular weight DNA.

In summary, we have identified a total of 19 novel haplotypes in the 3q29 region of the human genome, some of which were shown to have significant enrichment in certain populations. We have further shown that the majority of patients with the 3q29 deletion or duplication syndromes have breakpoints within a ∼5 kbp (blue) SD segment but find no evidence of inversions or other SVs to act as predisposing factors. Haplotypes identified in both 3q29 patients and unaffected individuals will be a valuable resource for future 3q29 deletion and duplication studies both to help to identify the breakpoints more precisely and whether certain 3q29 haplotypes predispose or protect from 3q29 deletion or duplication syndrome.

## Data Availability

All data produced in the present study are available upon reasonable request to the authors

## Supplemental Data

**Figure S1:**
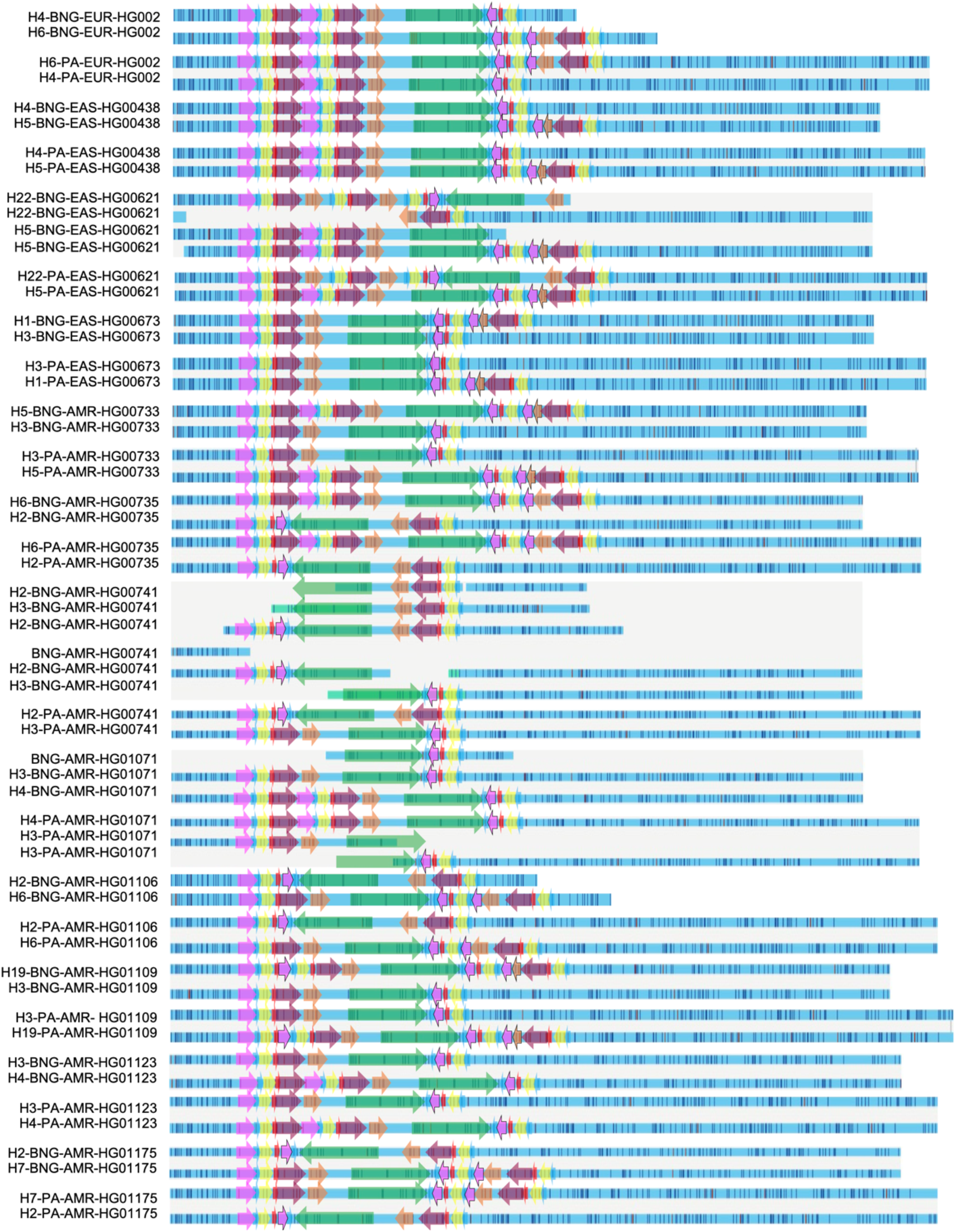
HPRC optical mapping and phased assembly 3q29 haplotype comparison – Part 1. EUR: European ancestry, EAS: East Asian ancestry, AMR: American ancestry, BNG: Bionano Genomics optical mapping data, PA: Phased assembly in silico mapping data, colored arrows depicting the 3q29 segments.

**Figure S2:**
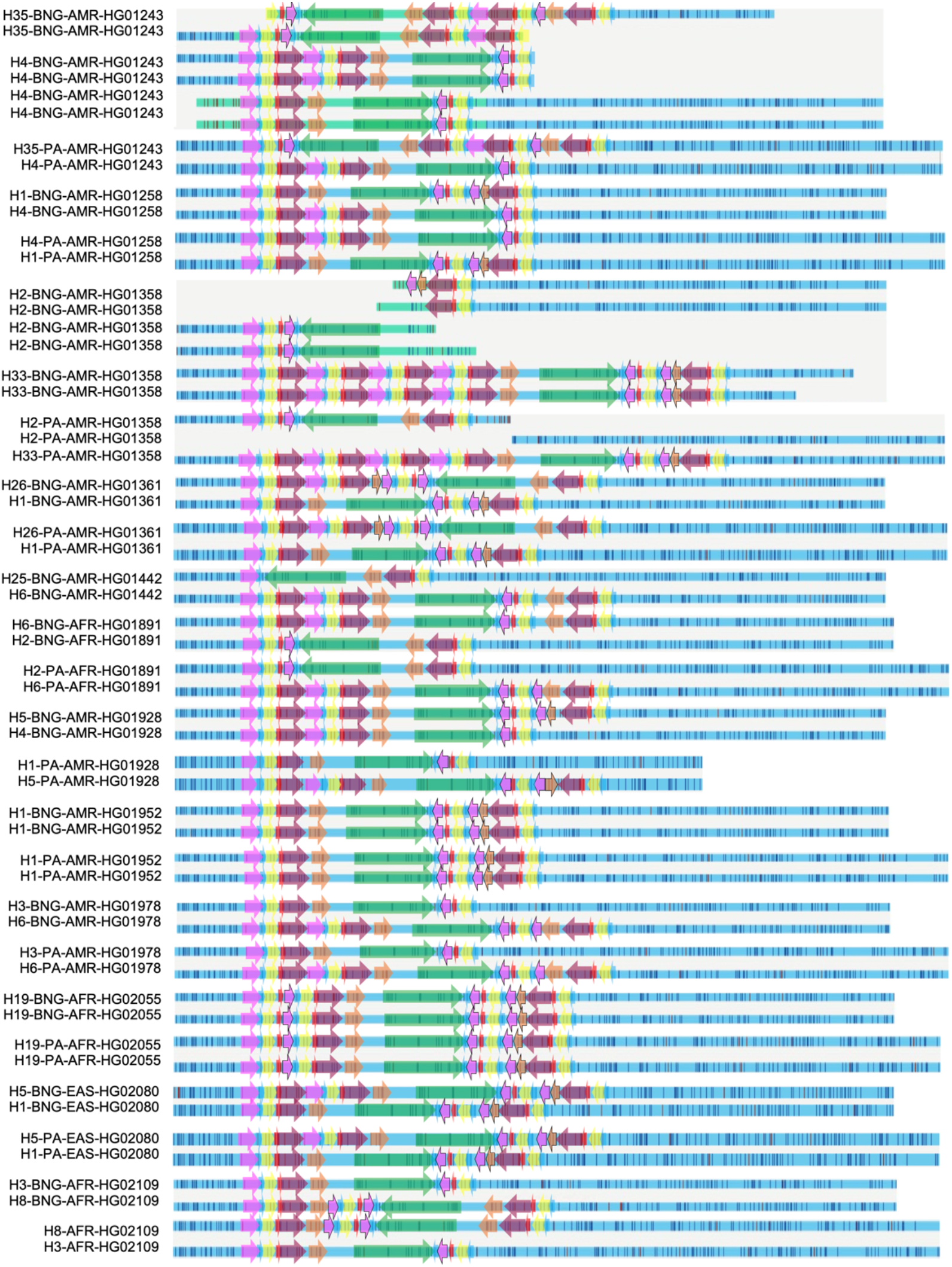
HPRC optical mapping and phased assembly 3q29 haplotype comparison – Part 2. AMR: American ancestry, EAS: East Asian ancestry, AFR: African ancestry, BNG: Bionano Genomics optical mapping data, PA: Phased assembly in silico mapping data, colored arrows depicting the 3q29 segments.

**Figure S3:**
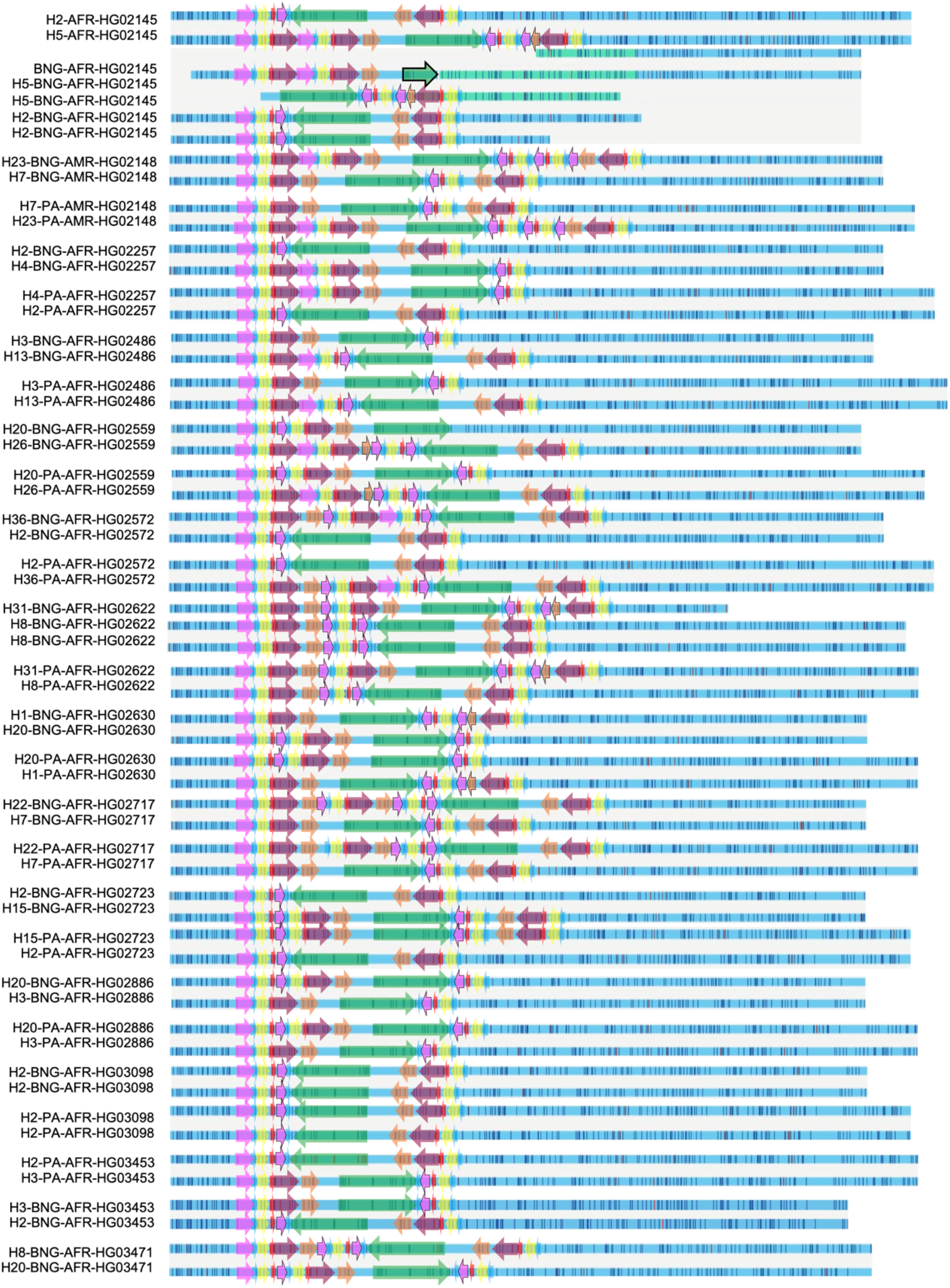
HPRC optical mapping and phased assembly 3q29 haplotype comparison – Part 3. AFR: African ancestry, AMR: American ancestry, BNG: Bionano Genomics optical mapping data, PA: Phased assembly in silico mapping data, colored arrows depicting the 3q29 segments.

**Figure S4:**
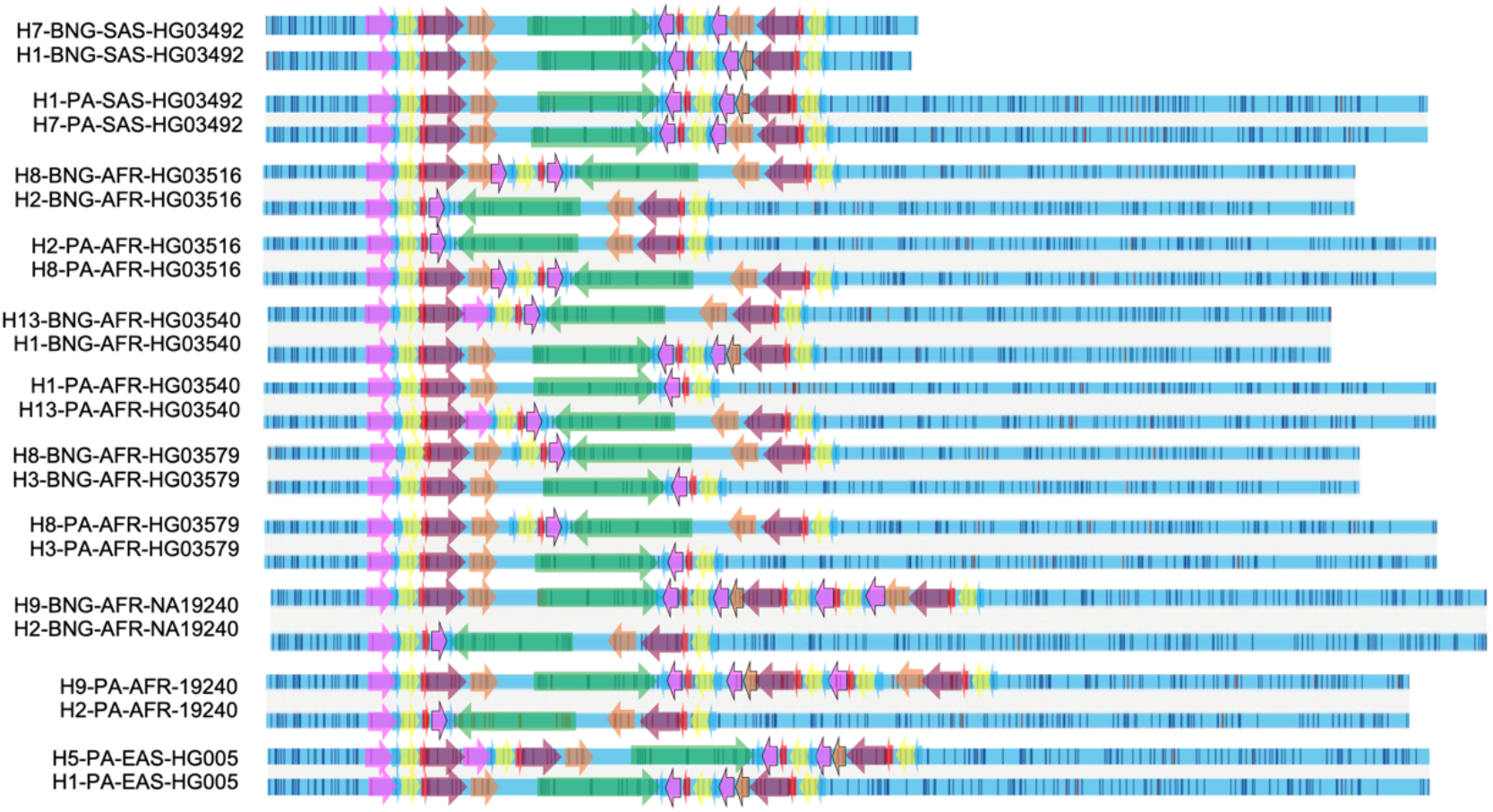
HPRC optical mapping and phased assembly 3q29 haplotype comparison – Part 3. AFR: African ancestry, AMR: American ancestry, BNG: Bionano Genomics optical mapping data, PA: Phased assembly in silico mapping data, colored arrows depicting the 3q29 segments.

**Figure S5:**
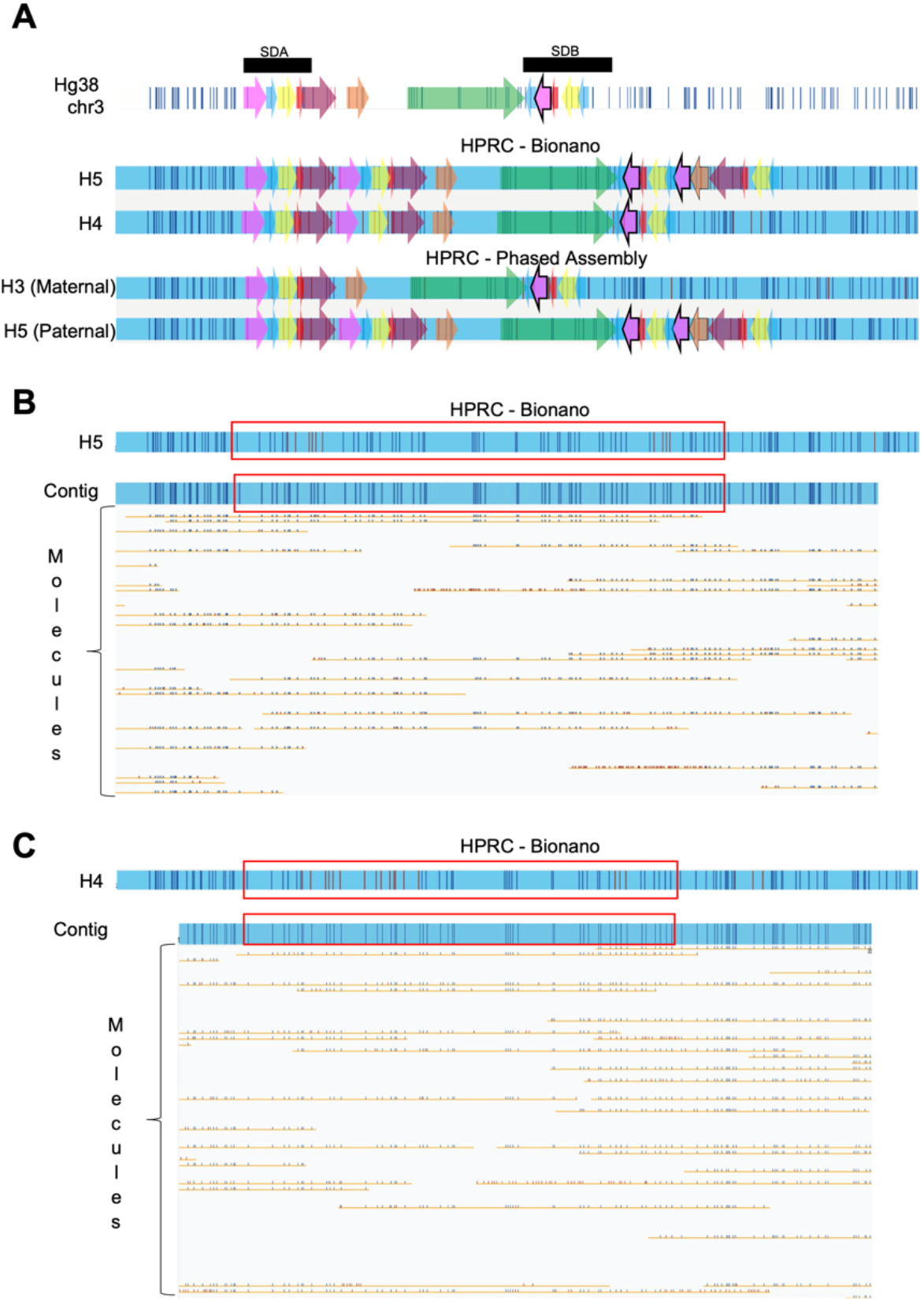
HPRC HG01928 bionano optical mapping and phased assembly 3q29 haplotype comparison. A – Hg38/GRCh38 in silico map represented at the top, optical mapping haplotypes, H5 and H4 respectively, represented in the middle, and phased assembly maternal and paternal haplotypes are represented at the bottom. B - Optical mapping of the H5 haplotype contigs represented in blue and optical mapping molecules represented as yellow lines showing single molecule support for the H5 haplotype. C - Optical mapping of the H4 haplotype contigs represented in blue and optical mapping molecules represented as yellow lines showing single molecule support for the H4 haplotype. The red boxes in B and C shows the 3q29 region in each haplotype.

**Figure S6:**
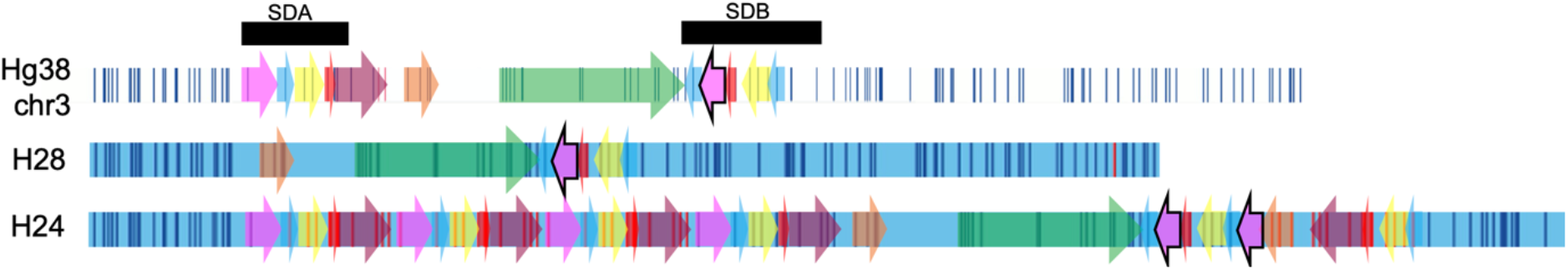
Comparison of the smallest, H28 (∼287 kbp), and the largest, H24 (∼859 kbp), 3q29 haplotypes. Colored blocks represent 3q29 segments, white rectangles with vertical lines represent Hg38/GRCh38 in silico map, and blue rectangles with vertical lines represent H28 and H24 haplotypes.

**Figure S7:**
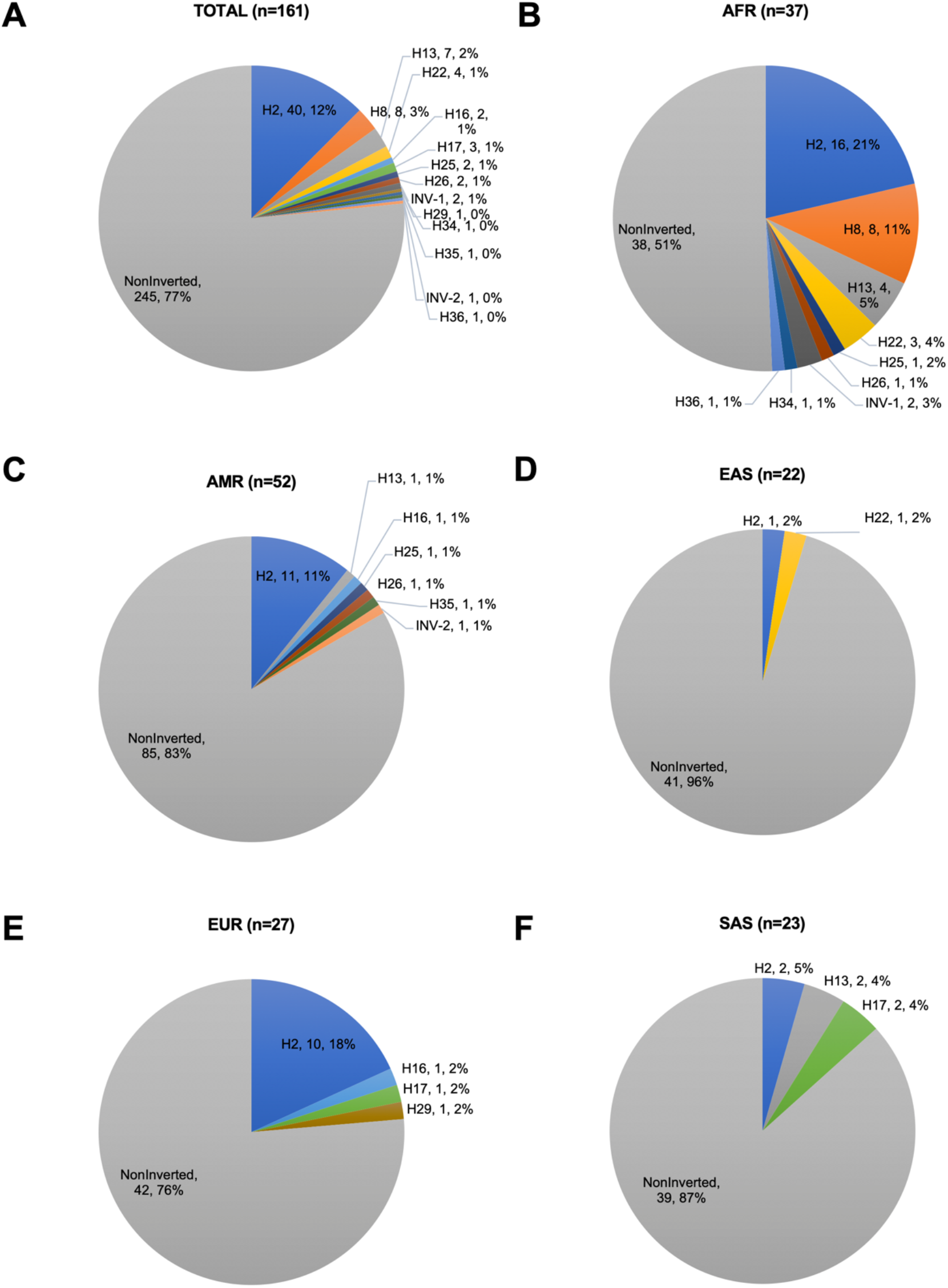
Inversion Frequencies: Inversions identified in this study; INV-1 and INV-2, and 289 kbp inversion. A - The proportion of inversion haplotypes among all studied samples, B - The proportion of inversion haplotypes in AFR population, C - The proportion of inversion haplotypes in AMR population, D - The proportion of inversion haplotypes in EAS population, E - The proportion of inversion haplotypes in EUR population, F - The proportion of inversion haplotypes in SAS population. Proportions were represented as: Haplotype ID/Count/Percentage.

**Figure S8:**
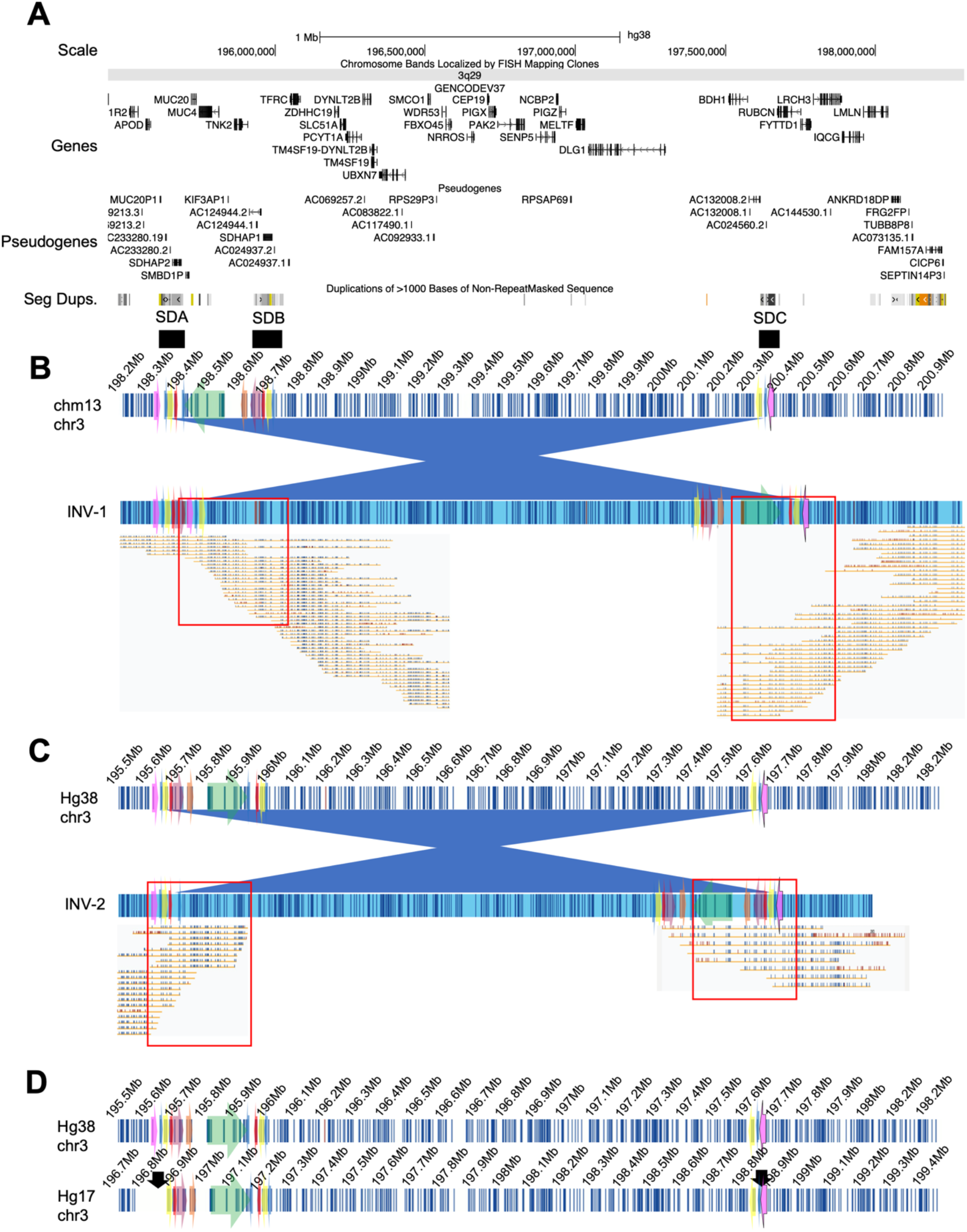
Large inversions identified in this study; INV-1 (∼2.03 Mbp) and INV-2 (∼2.13 Mbp). A - UCSC Genome Browser Genes, Pseudogenes, and segmental duplication tracks are represented. B - Upper rectangle with white background and vertical lines represents chm13/T2T (Telomere-to-Telomere) in silico map of the 3q29 region. The rectangle at the bottom represents the structure of INV-1. Red rectangle: highlighting the molecules supporting the inversion breakpoints. C - Upper rectangle with white background and vertical lines represents GRCh38/hg38 in silico map of the 3q29 region. The rectangle at the bottom represents the structure of INV-2. Red rectangle: highlighting the molecules supporting the inversion breakpoints. D - GRCh38/hg38 and NCBI35/hg17 reference assemblies represented with white backgrounds and vertical blue lines. Black arrows in NCBI35/hg17 indicate inversion breakpoints identified previously by Antonacci and colleagues in 2009. Colored arrows in each panel represent 3q29 segments.

**Figure S9:**
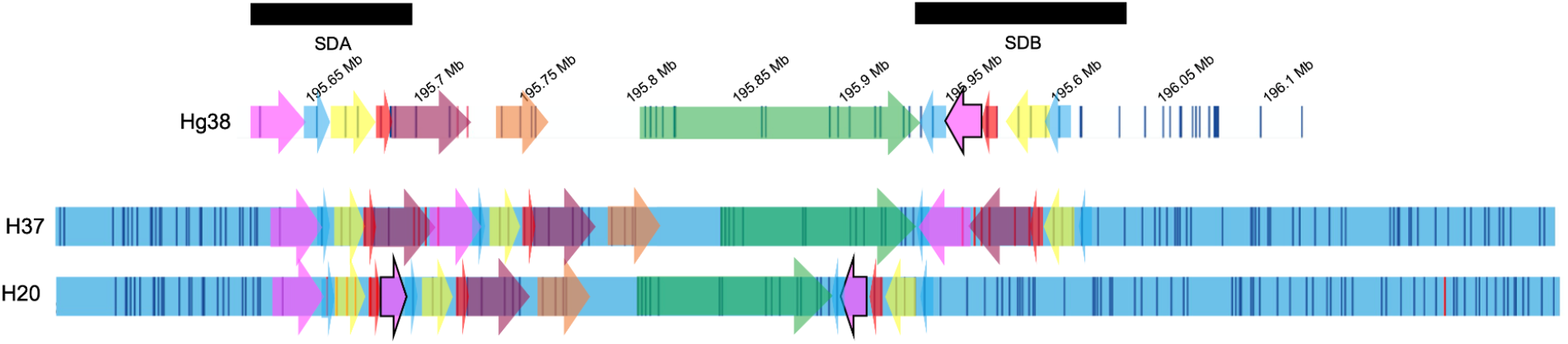
Novel haplotype H37 identified in one of the parents of origin (Father, Family 18) from the 3q29 Project. Optical mapping contigs in blue rectangles with vertical dark lines. Colored arrows represent the 3q29 segments in each haplotype.

Table S1: 1000 Genomes Project and California Initiative to Advance Precision Medicine Sample Cohort Optical Mapping Statistics

Table S2: The Emory 3q29 Project Sample Cohort Optical Mapping Statistics

Table S3: The 3q29 Segments

Table S4: Segment Sequence Identity

Table S5: 1000 Genomes Project and California Initiative to Advance Precision Medicine Sample Cohort Haplotypes

Table S6: DGV Overlap

Table S7: The Emory 3q29 Project Sample Cohort Haplotypes

Table S8: Breakpoint classes identified in the 3q29 Project probands with the 3q29 deletion or duplication syndrome

Table S9: The Emory 3q29 Project Proband Breakpoint Classes and Phenotype Comparison

## Declaration of Interests

The authors declare no competing interests.

## Acknowledgments

The authors are grateful to the participants and their families, without whose support this work would not have been possible. In addition, we thank the Human Genome Structural Variation Consortium and the Human Pangenome Reference Consortium for making their datasets available. We thank Dr. Pille Hallast, Dr. Jee Young Kwon, and Dr. Kwondo Kim for providing valuable feedback in editing this manuscript and Dr. Joshy George for assistance in statistical analyses, and The Jackson Laboratory High-Performance Computing admins for providing an environment to perform all computational analyses. J.G.M. conceived the study and analysis using Bionano optical mapping. F.Y. and U.G. performed the analysis and interpretation. J.G.M. obtained informed consent from parents and patients during hospital follow-up and facilitated collection of blood samples. J.G.M. coordinated study and subject enrollment. J.G.M. and T.M. carried out sample processing and experimentation. F.Y., and U.G drafted, and F.Y., C.L. and J.G.M. critically revised the article. Final approval of the version to be published was given by F.Y., U.G., T.M, Y.M., T.H.S, M.E.Z., P-Y.K., C.L. and J.G.M.

## Funding

C.L. was supported by the National Institute of Health (NIH) U24HG007497 and The First Affiliated Hospital of Xi’an Jiaotong University. T.H.S. and P-Y.K. were supported by grant # GM120772, from the National Institute of General Medical Sciences of the National Institutes of Health (NIH). Human Pangenome Reference Consortium support comes from NHGRI: A Human Genome Reference Center (HGRC; <u>RFA-HG-19-004</u>), High-Quality Human Reference Genomes (HGRQ; <u>RFA-HG-19-002</u>), Genome Reference Representations (GRR; <u>RFA-HG-19-003</u>), and Technology development for complete sequencing of genomes (<u>NOT-HG-19-011</u>). JGM was supported by R01 MH110701.

## Web resources

SV Genotyping: https://github.com/yuliamostovoy/OMGenSV

Bionano Solve 3.5.1: https://bionanogenomics.com/support/software-downloads/

OMTools: https://github.com/TF-Chan-Lab/OMTools

The Human Genome Structural Variation Consortium dataset:

http://ftp.1000genomes.ebi.ac.uk/vol1/ftp/data_collections/HGSVC2/working/20200117_Bionano_optical_maps/

University of California San Francisco dataset: Bianano bnx files of 114 samples (including 52 1000GP samples and 65 CIAPM samples) were available under the following accession number PRJNA588278 in the NCBI BioProject database.

The Human Pangenome Reference Consortium dataset: https://github.com/human-pangenomics/HPP_Year1_Data_Freeze_v1.0

## Data and code availability

The 3q29 Project dataset: The data supporting the findings of this study are available upon request from the authors.

HGSVC samples molecule support: 10.6084/m9.figshare.16899313

UCSF samples molecule support: 10.6084/m9.figshare.16886500

## Notes

### Competing Interest Statement

The authors have declared no competing interest.

### Funding Statement

C.L. was supported by the National Institute of Health (NIH) U24HG007497 and The First Affiliated Hospital of Xian Jiaotong University. T.H.S. and P-Y.K. were supported by grant #GM120772 from the National Institute of General Medical Sciences of the National Institutes of Health (NIH). Human Pangenome Reference Consortium support comes from NHGRI: A Human Genome Reference Center (HGRC- RFA-HG-19-004) High-Quality Human Reference Genomes (HGRQ- RFA-HG-19-002) Genome Reference Representations (GRR- RFA-HG-19-003) and Technology development for complete sequencing of genomes (NOT-HG-19-011). JGM was supported by R01 MH110701.

### Author Declarations

This study was approved by Emory University IRB (IRB #000088012).

### Summary of Updates

The title of the manuscript is updated.

